# Association between Hormonal Contraceptive Use and Lipedema: A Cross-Sectional Study with 637 Brazilian Women

**DOI:** 10.64898/2025.12.05.25341470

**Authors:** Alexandre Campos Moraes Amato, Juliana Lelis Spirandeli Amato, Daniel Augusto Benitti

## Abstract

**Background:** Lipedema is a chronic condition characterized by disproportionate adipose tissue accumulation, pain, and sensitivity, often influenced by hormonal fluctuations.

Despite its prevalence, the specific impact of exogenous hormones on the disease course remains understudied.

**Objective:** To investigate the association between hormonal contraceptive use and the presence and severity of lipedema in Brazilian women.

**Methods:** A cross-sectional study was conducted at Amato – Instituto de Medicina Avançada using a structured online questionnaire administered between August and November 2025.

Brazilian women aged 18 years or older with suspected or diagnosed lipedema were included. The questionnaire assessed demographic and clinical characteristics, history of contraceptive use, lipedema symptoms, and impact on quality of life. Symptom scores (0-8 points) and quality of life impact scores (0-15 points) were calculated. Statistical analyses included Chi-square and Kruskal-Wallis tests, Spearman correlations, and logistic and linear regressions.

**Results:** A total of 637 women participated, with a mean age of 41.8 ± 8.7 years and a mean BMI of 28.9 ± 6.4 kg/m². Of the participants, 491 (77.1%) had a confirmed diagnosis of lipedema. It was observed that 58.8% of users reported symptom worsening after contraceptive use (p<0.001). Free-text analysis revealed that 15.1% of participants experienced the onset of lipedema symptoms concurrently with the start of contraceptive use. Weight gain as a side effect was strongly associated with worsening. In multivariate logistic regression, symptom score (OR=1.562, p<0.001) and age at menarche (OR=0.746, p=0.0135) were significant predictors of worsening. Regarding impact on quality of life, BMI (beta=0.364) and pain (beta=0.641) were independent predictors. Conclusions: This study demonstrates a significant association between hormonal contraceptive use and self-reported worsening of lipedema symptoms. These results have potential implications for individualized contraceptive counseling for women with lipedema and reinforce the need for prospective investigations to confirm the nature and direction of this association.

## INTRODUCTION

Lipedema is a chronic and progressive disease of adipose tissue that predominantly affects women, characterized by symmetrical and painful fat accumulation in the limbs, associated with edema, increased sensitivity, and resistance to weight loss [1,2]. With an estimated prevalence of 12.3% in the female population, it represents a significant impact on the quality of life and functionality of affected women [3].

The etiology of lipedema remains incompletely elucidated. Evidence suggests a genetic component and a modulating role of female sex hormones, as the disease frequently manifests or worsens during periods of hormonal fluctuation (puberty, pregnancy, menopause). Particularly, hormonal contraceptives (HC) have been anecdotally associated with symptom worsening, although robust scientific evidence is scarce [4,5].

The biological plausibility of this association lies in multiple mechanisms. The estrogenic components of HCs may amplify mast cell activation, alter vascular permeability, compromise lymphatic function, and influence adipocyte proliferation. Additionally, common side effects such as weight gain and fluid retention may exacerbate clinical manifestations [6,7].

In Brazil, where HC use is prevalent and lipedema remains underdiagnosed, understanding this relationship is of extreme clinical and public health relevance [8]. This cross-sectional study aimed to investigate the association between hormonal contraceptive use and the presence, severity, and progression of lipedema in a sample of 637 Brazilian women, utilizing an innovative methodological approach with natural language processing (NLP) via Large Language Models (LLM) for qualitative data enrichment. We sought to quantify this association, identify predictors of worsening, explore intermediate mechanisms, and assess the impact on quality of life, providing evidence for safer contraceptive counseling in this vulnerable population.

## METHODS

### Study Design

This was an observational cross-sectional study based on a self-administered online questionnaire, conducted at Amato – Instituto de Medicina Avançada between August and November 2025.

### Population and Sample

The study included women aged 18 years or older residing in Brazil with suspected or confirmed lipedema who consented to participate. Incomplete questionnaires (with less than 50% of questions answered), duplicates identified by IP address, and inconsistent or biologically implausible data were excluded.

Participant recruitment was carried out through dissemination on social media (Instagram, Facebook, WhatsApp), support groups for women with lipedema, specialized clinics and offices, and patient associations. The final sample consisted of 637 participants.

### Data Collection Instrument

A structured questionnaire developed specifically for this study was used, consisting of eight sections. Section A collected demographic data (age, weight, height, geographic location) and the informed consent form. Section B comprised nine lipedema screening questions, investigating leg pain, progressive swelling, worsening with heat, sensitivity to touch, easy bruising, disproportionate fat distribution, resistance to diet, family history, and timing of symptom onset.

Section C assessed the diagnosis and clinical characteristics of lipedema, including medical diagnosis confirmation, date of diagnosis, specialty of the responsible physician, lipedema Stage (1 to 4), lipedema Type (I to IV), and associated comorbidities. Section D collected information on hormonal and reproductive history, covering age at menarche, menstrual cycle regularity, number of pregnancies and miscarriages, menopausal status, use of hormone replacement therapy, and thyroid alterations.

Section E, the main focus of this study, consisted of 14 questions regarding contraceptive use, including current use status, type of contraceptive (pill, IUD, implant, vaginal ring, injection), commercial name of the product, current and total duration of use, age at onset of use, pattern of use (continuous versus interrupted), reasons for discontinuation, perceived side effects, perception of lipedema worsening, changes in symptoms, associated weight gain, and alternative contraceptive methods used.

Section F assessed the impact on quality of life through questions on mobility, difficulties in daily activities, impact on work, social and emotional impact, visual analog scale for pain (0–10), sleep quality, self-esteem, and perception of family support. Section G investigated current treatments (lymphatic drainage, compression stockings, physical therapy, nutritional and psychological follow-up, lipedema surgery) and satisfaction with treatments. Finally, Section H allowed for additional observations in a free-text field and verified participants’ interest in receiving the results.

The complete structure of the questionnaire, translated into English, is provided in Appendix Table 1.

### Study Variables

The primary outcome variable was lipedema worsening associated with hormonal contraceptive use, categorized as "No difference noticed," "Yes, slightly," "Yes, severe worsening," and "Improved," from which a binary variable (worsening yes/no) was also derived. Secondary outcomes considered were the lipedema symptom score (0–8 points), the quality-of-life impact score (0–15 points), the pain scale (0–10 points), and the self-esteem level (Low, Fair, Good, Excellent).

The main explanatory variables included contraceptive use status (current, previous, never), the type of contraceptive used, the duration of use in years, and the age at onset of use.

Control variables (covariates) included age in years, Body Mass Index (BMI in kg/m²), age at menarche, time since menarche, number of comorbidities, Lipedema Stage, and current treatments.

### Data Processing and Enrichment

Raw data underwent multi-step processing. Initially, cleaning and validation were performed, including duplicate removal, consistency checks, and treatment of missing values. Subsequently, derived variables were calculated: BMI using the formula weight (kg) divided by height squared (m²), BMI categories according to WHO criteria [9], five-year age groups, symptom score (sum of "Yes" responses to the eight questions in Section B), symptom severity (Mild, Moderate, Severe, Very Severe), quality of life impact score (weighted sum of the five questions in Section F), duration of contraceptive use extracted from free text, and years since menarche.

Additionally, data enrichment was performed using natural language processing (NLP) via a local language model (LM Studio, model openai/gpt-oss–20b). Free-text fields were processed for the automatic categorization of side effects into eight classes, including weight gain, mood swings, headache, nausea, reduced libido, swelling, bleeding, and others. Furthermore, the model performed the extraction and classification of treatments into eight modalities, such as diet, exercise, compression stockings, lymphatic drainage, medications, surgery, and physical therapy. The processing also included the identification and standardization of associated comorbidities and the temporal classification of symptom onset into categories such as puberty, pregnancy, menopause, and contraceptive use. A random sample (e.g., 10%) of the automated categorizations was manually reviewed to ensure consistency and accuracy

## Statistical Analysis

Statistical analyses were performed using Python 3.11 with the pandas, numpy, scipy, statsmodels, and scikit-learn packages. For descriptive analysis, continuous variables were presented as mean ± standard deviation and median with interquartile range (IQR), while categorical variables were presented as absolute and relative frequencies. The normality of distributions was assessed using the Shapiro-Wilk test.

In inferential analysis, bivariate tests were employed, including the Chi-square test for associations between categorical variables, Student’s t-test or Mann-Whitney U test for comparing two groups, ANOVA or Kruskal-Wallis test for comparing three or more groups, and Spearman’s correlation for associations between continuous variables.

For multivariate analysis, a logistic regression model was adjusted to identify factors associated with worsening due to contraceptives. The dependent variable was worsening (binary), and independent variables included age, BMI, age at menarche, duration of use, symptom score, and number of comorbidities. Results were presented as Odds Ratios (OR) with 95% confidence intervals. Additionally, a multiple linear regression model was adjusted for factors associated with the impact on quality of life, with the quality-of-life impact score as the dependent variable and age, BMI, symptom score, pain scale, degree of worsening with contraceptives, and number of comorbidities as independent variables.

Results were presented as standardized β coefficients.

A significance level of 5% (α = 0.05, two-tailed) was adopted for all statistical tests. P-values were adjusted for multiple comparisons when appropriate using the Bonferroni method.

### Ethical Aspects

This study was conducted in accordance with the ethical principles of the Declaration of Helsinki and Brazilian resolutions (CNS 466/2012 and 510/2016). All participants provided electronic consent after reading the Informed Consent Form. Data were anonymized and stored securely with AES-256 encryption. Ethics Committee Approval: CAAE 90936425.0.0000.0081.

## DATA AVAILABILITY

The anonymized data and the code for reproducing the analyses are available from the authors upon reasonable request, in compliance with data protection guidelines (LGPD).

## RESULTS

A total of 637 women participated in this study, representing the largest Brazilian sample investigated on lipedema to date. The demographic and clinical characteristics revealed a mean age of 41.8 ± 8.7 years (range: 19-68 years) and a mean BMI of 28.9 ± 6.4 kg/m², with the majority of participants (77.1%, n=491) having a confirmed diagnosis of lipedema by a healthcare professional. The geographic distribution showed predominance of participants from the Southeast region (64.8%), followed by the South (16.6%), which reflects both the regional concentration of specialized lipedema care centers and the demographic distribution of the Brazilian population. Detailed demographic and clinical characteristics of the sample are presented in Table 1.

**Table 1 -.** Demographic and clinical characteristics of study participants (n=637)

| Variable | Value |
| --- | --- |
| **Age (years)** |  |
| Mean ± SD | 41.8 ± 8.7 |
| Median [IQR] | 42.0 [35.0–47.0] |
| Min-Max | 18–73 |
| **BMI (kg/m<sup>2</sup>)** |  |
| Mean ± SD | 28.9 ± 6.4 |
| Median [IQR] | 27.7 [24.8–32.0] |

SD = standard deviation; IQR = interquartile range; BMI = body mass index. Data are presented as mean ± SD, median [IQR], and range (min-max) for continuous variables, and as n (%) for categorical variables.

Regarding the clinical characteristics of lipedema, the distribution across disease stages revealed that Stage 2 was the most prevalent (51.8%, n=254), followed by Stage 3 (26.3%, n=129) and Stage 1 (19.0%, n=93), with only a small proportion in Stage 4 (2.9%, n=14). The most common lipedema type was Type III, affecting hips, thighs, and calves (58.1%, n=285), followed by Type II involving hips and thighs (32.9%, n=161). A substantial proportion of participants reported comorbidities, with hypothyroidism being the most frequent (28.1%, n=179), followed by anxiety/depression (22.6%, n=144) and hypertension (18.5%, n=118). The burden of associated conditions was considerable, with a median of 2 comorbidities per participant. The clinical characteristics of lipedema in our sample are detailed in Table 2.

**Table 2 -.** Clinical characteristics and disease severity of lipedema (n=637)

| Characteristic | n (%) |
| --- | --- |
| **Confirmed Diagnosis** |  |
| Yes | 491 (77.1%) |
| Suspected | 131 (20.6%) |
| No | 15 (2.4%) |
| **Lipedema Stage** |  |
| Stage 2 | 205 (32.2%) |
| Stage 1 | 113 (17.7%) |
| Stage 3 | 106 (16.6%) |
| Do not know | 79 (12.4%) |
| Stage 4 | 21 (3.3%) |
| **Symptom Score (0–8)** |  |
| Mean ± SD | 6.07 ± 1.54 |

Data are presented as n (%) for categorical variables and mean ± SD for continuous variables. Percentages may not total 100% due to missing data or rounding.

The pattern of hormonal contraceptive use in the study population demonstrated that 588 participants (92.3%) had current or previous experience with hormonal contraception.

Among those with contraceptive experience, oral contraceptive pills were by far the most commonly used method (88.9%, n=523), followed by hormonal IUDs (18.7%, n=110) and injectable contraceptives (15.0%, n=88). The median duration of use was 7.0 years (IQR: 3.0-15.0 years), with a mean of 10.3 ± 9.1 years, indicating substantial long-term exposure to exogenous hormones in this population. The age at onset of contraceptive use averaged 20.7 ± 5.6 years, typically coinciding with the period of reproductive maturity. Among current users, continuous use patterns predominated (67.4%), while 32.6% reported interrupted use with periods of discontinuation. The comprehensive pattern of hormonal contraceptive use is presented in Table 3.

**Table 3 -.**
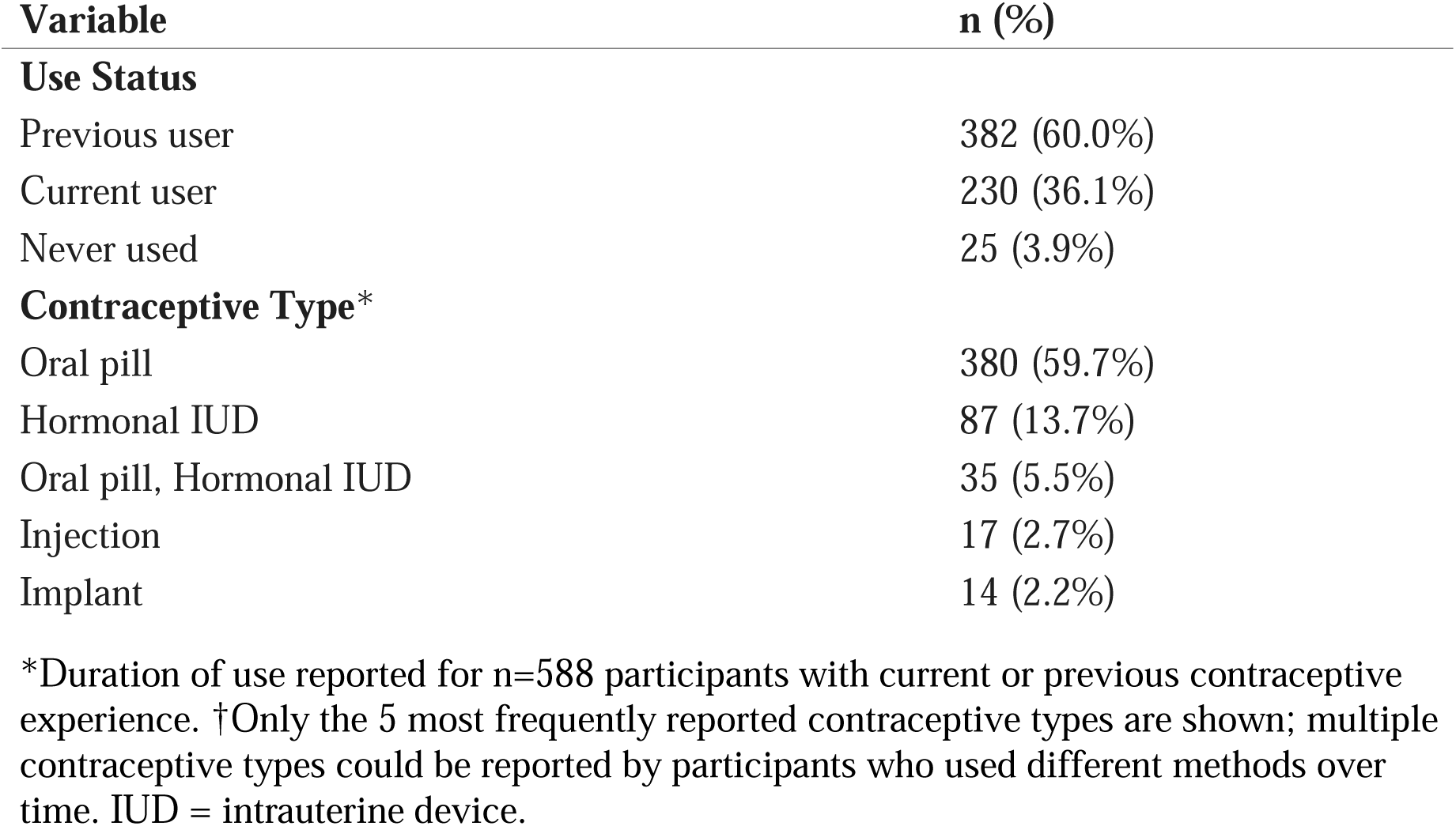
Pattern and type of hormonal contraceptive use among study participants (n=637)

| Variable | n (%) |
| --- | --- |
| **Use Status** |  |
| Previous user | 382 (60.0%) |
| Current user | 230 (36.1%) |
| Never used | 25 (3.9%) |
| **Contraceptive Type\*** |  |
| Oral pill | 380 (59.7%) |
| Hormonal IUD | 87 (13.7%) |
| Oral pill, Hormonal IUD | 35 (5.5%) |
| Injection | 17 (2.7%) |
| Implant | 14 (2.2%) |

\*Duration of use reported for n=588 participants with current or previous contraceptive experience. †Only the 5 most frequently reported contraceptive types are shown; multiple contraceptive types could be reported by participants who used different methods over time. IUD = intrauterine device.

One of the most clinically significant findings of this study was the substantial prevalence of lipedema symptom worsening associated with hormonal contraceptive use. Among the 588 participants who had used contraceptives, 346 (58.8%) reported experiencing some degree of symptom exacerbation. Specifically, 203 participants (34.5%) reported severe worsening, while 143 (24.3%) noted slight worsening. Only 237 participants (40.3%) perceived no difference in symptoms, and remarkably, only 5 participants (0.9%) reported improvement. This distribution of responses is detailed in Table 4 and visually represented in Figure 1. The statistical significance of this association was confirmed through Chi-square testing, which demonstrated that the distribution of responses differed markedly from the expected uniform distribution (χ² = 213.71, p < 0.001), definitively indicating that the perception of worsening was not a random phenomenon.

**Figure 1 -.**
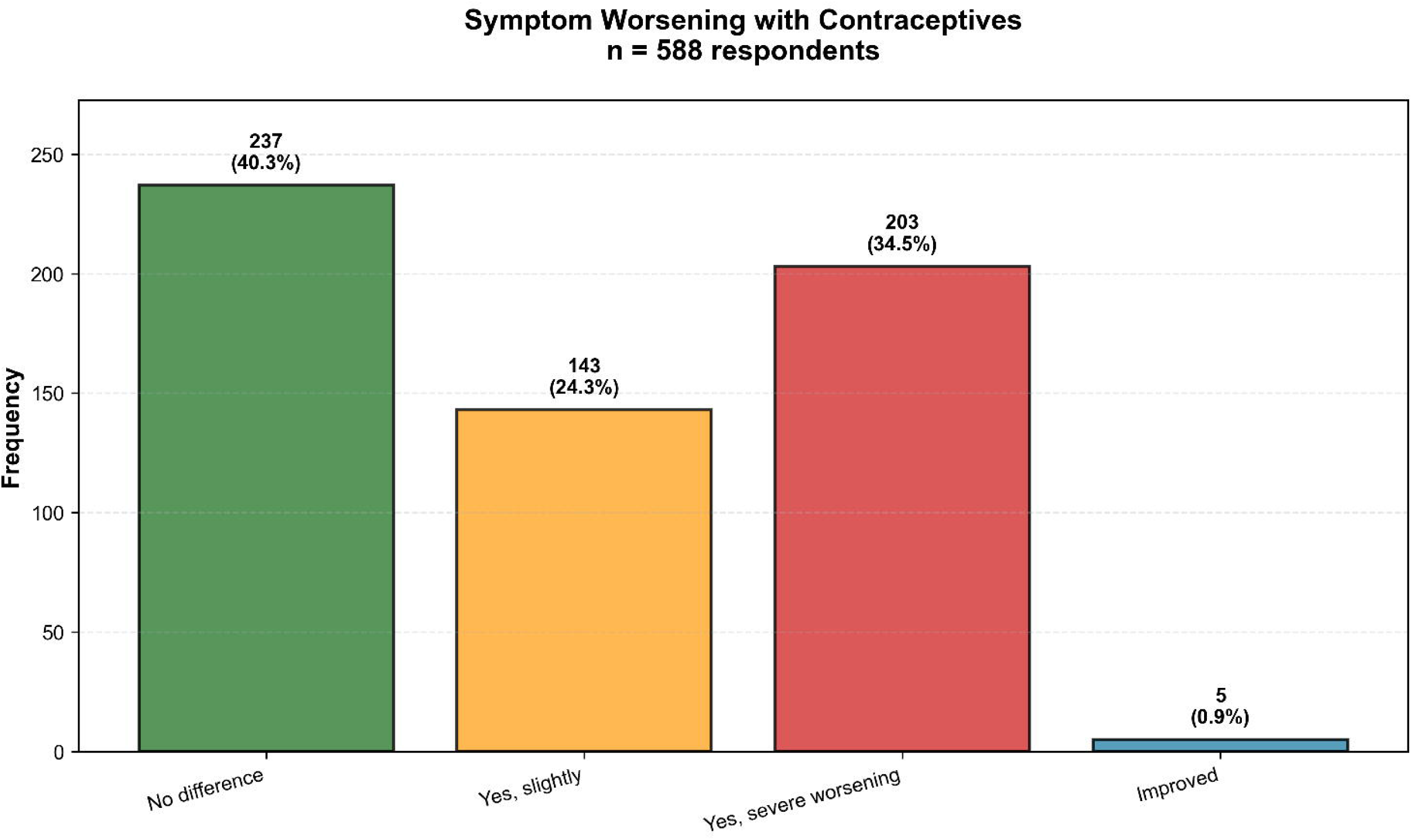
Distribution of perceived lipedema symptom changes with hormonal contraceptive use (n=588). The majority of participants (58.8%) reported some degree of worsening: 34.5% severe and 24.3% slight. Only 40.3% noticed no difference, and 0.9% reported improvement.

**Table 4 -.**
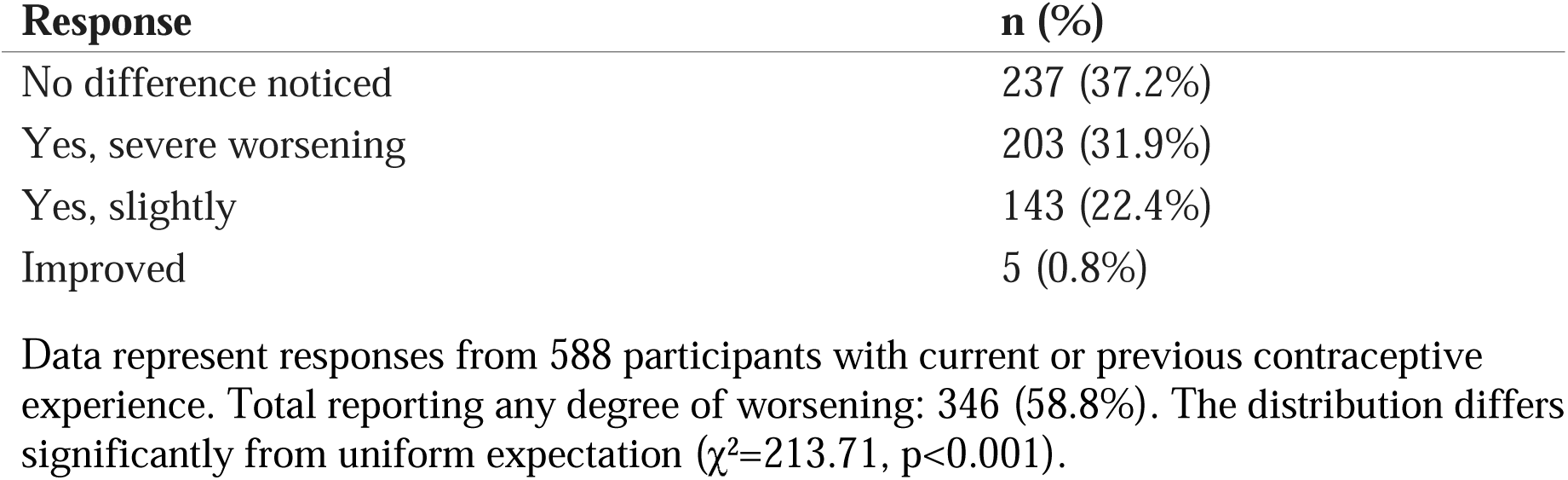
Distribution of perceived lipedema symptom changes associated with hormonal contraceptive use (n=588)

A particularly intriguing temporal finding emerged from the free-text analysis using natural language processing. When participants were asked about the timing of lipedema symptom onset, 15.1% (n=96) specifically reported that their symptoms began concurrently with the initiation of contraceptive use, suggesting a possible temporal association that warrants further prospective investigation. This finding is depicted in Figure 2, which shows that while puberty remained the most common period of symptom onset (39.7%, n=253), contraceptive use represented the third most frequently reported trigger period, following indeterminate timing (28.1%, n=179) and preceding pregnancy (11.6%, n=74) and menopause (5.5%, n=35).

**Figure 2 -.**
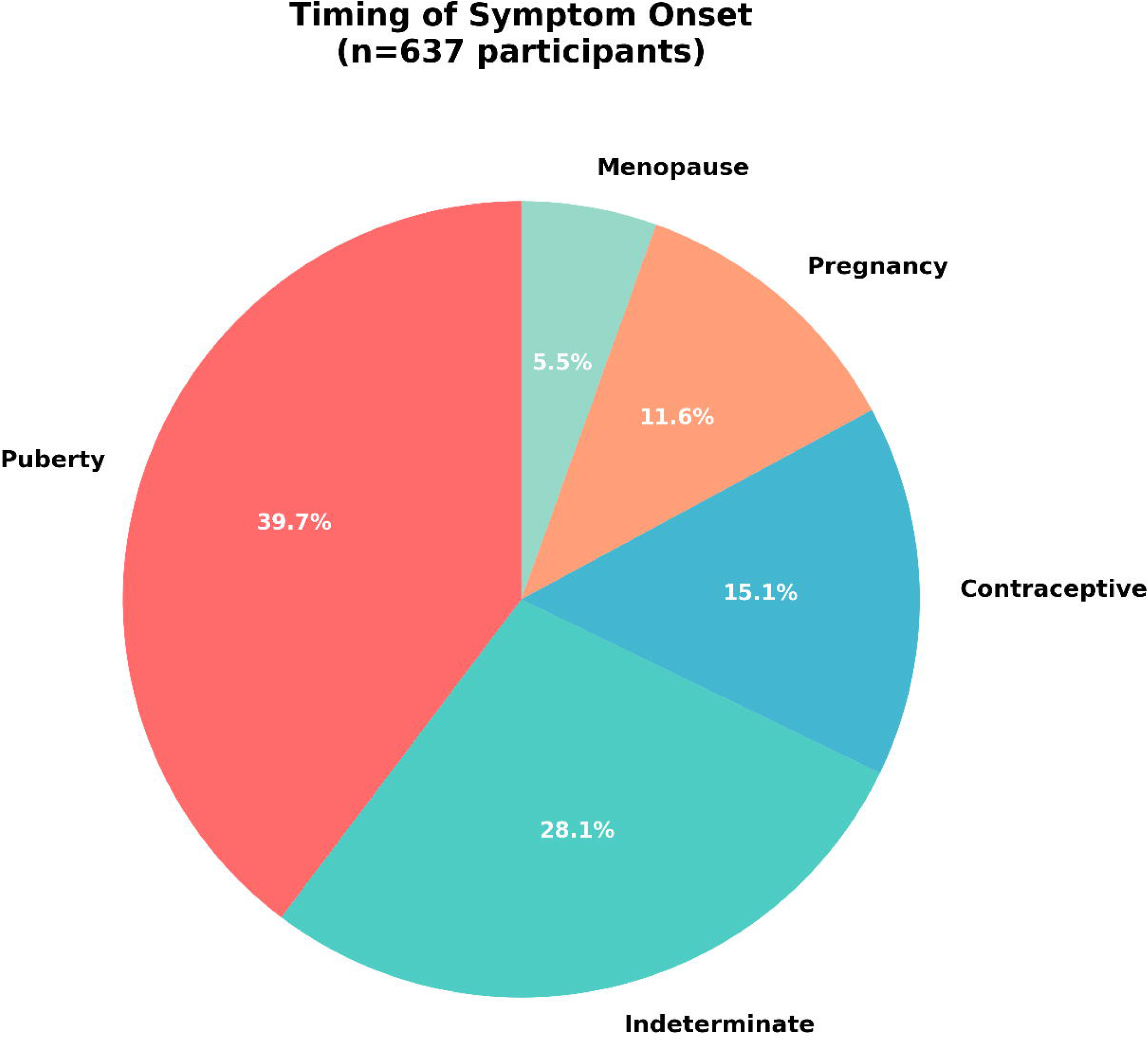
Temporal distribution of lipedema symptom onset (n=637). Puberty was the most common period of initial manifestation (39.7%), followed by indeterminate timing (28.1%). Contraceptive use (15.1%), pregnancy (11.6%), and menopause (5.5%) were also reported as trigger periods.

To better understand the mechanisms underlying contraceptive-associated worsening, we analyzed the relationship between specific side effects and symptom exacerbation. The most commonly reported side effects were weight gain (41.8%, n=266), swelling (40.5%, n=258), headache (22.0%, n=140), and mood changes (13.2%, n=84), as illustrated in Figure 3. Importantly, these side effects showed significant associations with lipedema worsening. Women who experienced weight gain as a contraceptive side effect were substantially more likely to report lipedema worsening (71.7%) compared to those without weight gain (43.5%), representing a clinically meaningful difference of 28.2 percentage points (χ² = 29.32, p < 0.0001). Similarly, mood alterations as a side effect were associated with higher rates of symptom worsening (77.4% versus 51.3% in those without mood changes, χ² = 14.23, p = 0.0002).

**Figure 3 -.**
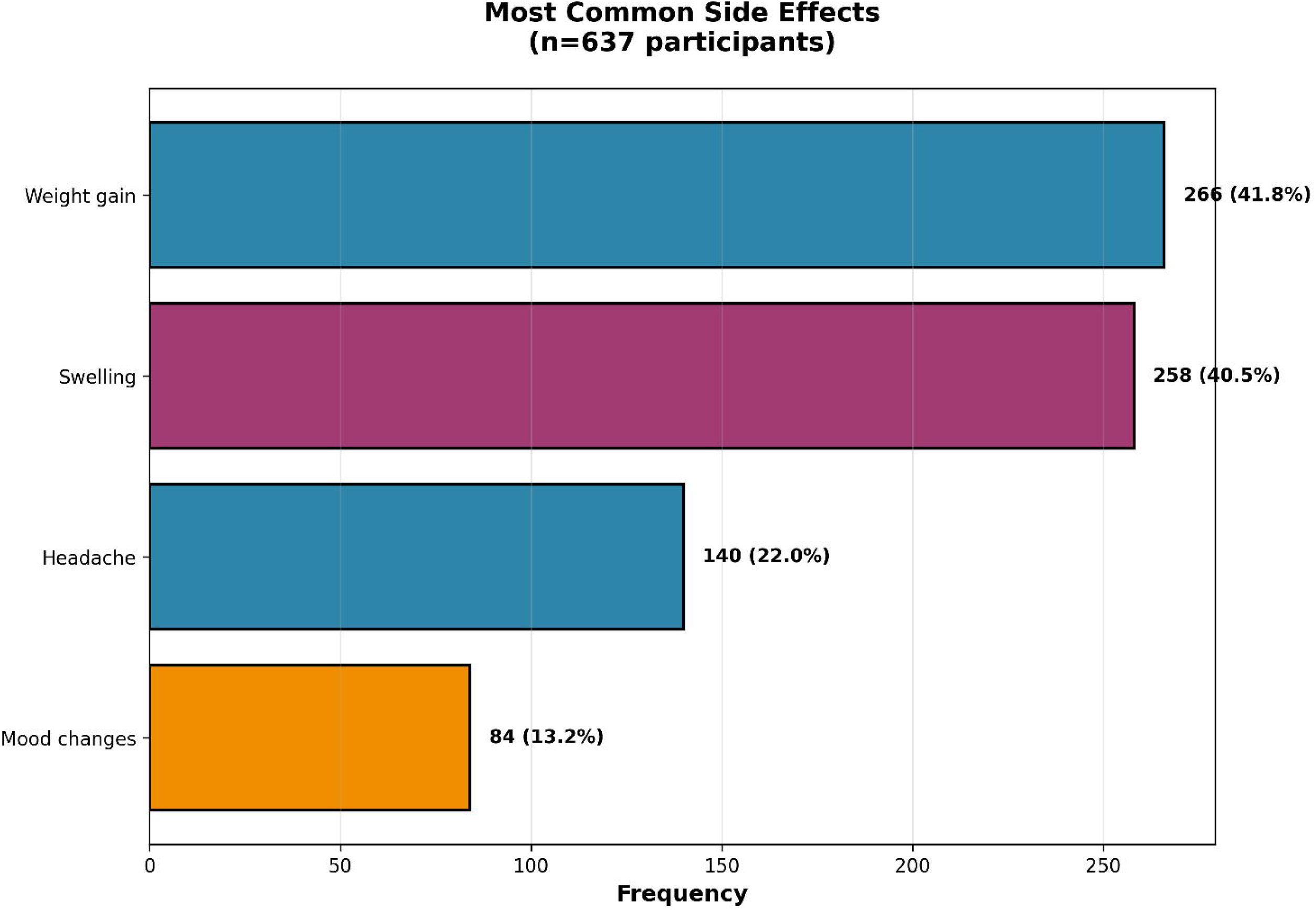
Prevalence of the most commonly reported side effects during hormonal contraceptive use (n=637). Weight gain (41.8%) and swelling (40.5%) were the most prevalent adverse effects, followed by headache (22.0%) and mood changes (13.2%).

The impact of lipedema on quality of life was substantial and objectively quantifiable. The pain scale (ranging from 0 to 10) showed a mean score of 5.2 ± 2.6, with a median of 5.0, indicating moderate to severe chronic pain in this population. The quality-of-life impact score (ranging from 0 to 15) demonstrated a mean of 3.09 ± 1.89, with a median of 3.0.

Self-esteem assessment revealed considerable psychosocial burden, with 70.5% of participants reporting either fair (38.3%, n=244) or low (32.2%, n=205) self-esteem, while only 22.3% reported good or excellent self-esteem. These findings are summarized in Table 5.

**Table 5 -.**
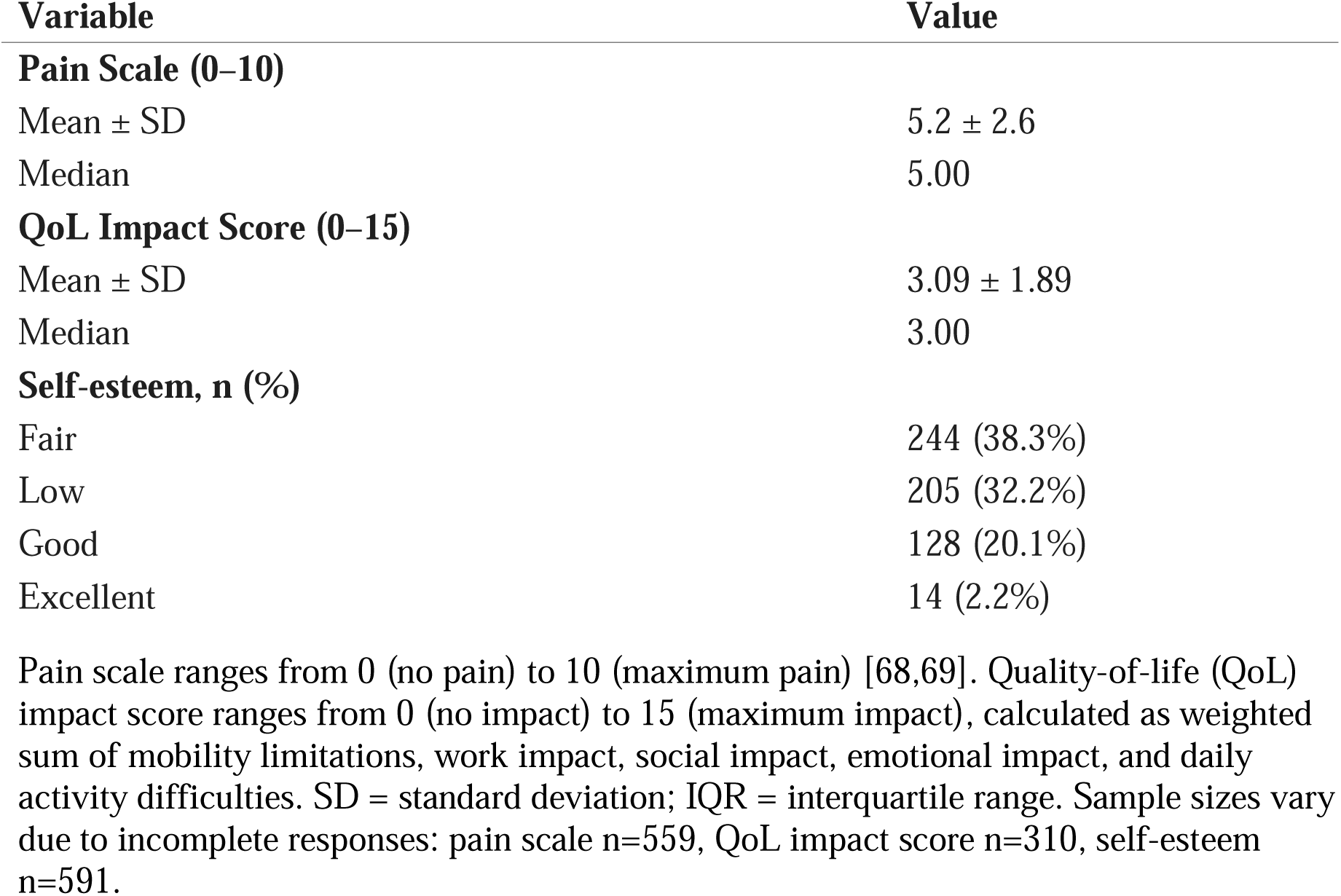
Pain intensity, quality-of-life impact, and self-esteem among study participants.

Correlation analyses revealed several statistically significant relationships among key variables (Table 6). As expected, age showed a very strong correlation with years since menarche (r=0.973, p<0.0001). More clinically relevant, BMI demonstrated a moderate positive correlation with quality-of-life impact score (r=0.313, p<0.0001), suggesting that higher body mass index is associated with greater functional impairment. The symptom score showed a moderate correlation with the pain scale (r=0.431, p<0.0001), and notably, the pain scale itself exhibited the strongest correlation with quality-of-life impact (r=0.500, p<0.0001), indicating that pain intensity is a primary driver of functional limitations in women with lipedema.

**Table 6 -.** Significant correlations among key clinical variables (Spearman rank correlation)

| Variable 1 | Variable 2 | r | p-value | n |
| --- | --- | --- | --- | --- |
| Age | Years since menarche | 0.973 | <0.0001 | 637 |
| BMI | QoL impact score | 0.313 | <0.0001 | 637 |
| Symptom score | Pain scale (F6) | 0.431 | <0.0001 | 559 |
| Pain scale (F6) | QoL impact score | 0.500 | <0.0001 | 559 |

Only correlations with $|r| > 0.3$ and $p < 0.05$ are shown. Correlation strength interpretation: $|r| = 0.3-0.5$ (moderate), $|r| = 0.5-0.7$ (strong), $|r| > 0.7$ (very strong). BMI = body mass index; QoL = quality of life; F6 = question F6 from study questionnaire (pain scale 0-10).

To identify independent predictors of lipedema worsening associated with contraceptive use while controlling for potential confounders, we conducted multivariate logistic regression analysis with 568 participants who had complete data. The model demonstrated modest overall fit (Pseudo R² = 0.048; AIC = 752.0; BIC = 782.4), suggesting that contraceptive-associated worsening is influenced by multiple factors beyond those measured in this study. The results, presented in Table 7, revealed that the symptom score was the strongest independent predictor of worsening (OR = 1.562; 95% CI: 1.300-1.877; p < 0.0001), indicating that each one-point increase in baseline symptom severity was associated with a 56% increase in the odds of experiencing contraceptive-related worsening. This finding suggests that women with more severe pre-existing symptoms are particularly vulnerable to hormonal exacerbation.

**Table 7 -.** Multivariate logistic regression model for predictors of lipedema worsening with contraceptive use (n=568)

| Variable | OR | 95% CI | p-value |
| --- | --- | --- | --- |
| Age | 0.926 | 0.776–1.105 | 0.3949 |
| BMI | 0.819* | 0.680–0.988 | 0.0369 |
| Age at menarche | 0.746* | 0.591–0.941 | 0.0135 |
| Duration of use (years) | 0.931 | 0.789–1.100 | 0.4018 |
| Symptom score | 1.562* | 1.300–1.877 | <0.0001 |
| Number of comorbidities | 0.904 | 0.759–1.076 | 0.2572 |

Dependent variable: any degree of symptom worsening (yes/no) during contraceptive use. All continuous variables were standardized prior to analysis. Model fit: Pseudo $R^2$ =0.048; AIC=752.0; BIC=782.4. \*p<0.05. OR = odds ratio; CI = confidence interval; BMI = body mass index. Sample includes only participants with complete data on all model variables.

Interestingly, two other variables showed significant but protective associations. Age at menarche demonstrated a protective effect (OR = 0.746; 95% CI: 0.591-0.941; p = 0.0135), suggesting that women with later onset of menstruation may have reduced susceptibility to contraceptive-associated worsening. BMI also showed a modest protective association (OR = 0.819; 95% CI: 0.680-0.988; p = 0.0369), a somewhat counterintuitive finding that may reflect selection bias or unmeasured confounding and warrants further investigation.

Notably, the duration of contraceptive use was not significantly associated with worsening (OR = 0.931; 95% CI: 0.789-1.100; p = 0.4018), suggesting that symptom exacerbation may be related to individual susceptibility rather than cumulative hormonal exposure.

Neither age nor number of comorbidities showed significant associations with worsening in the adjusted model.

To complement our understanding of the functional impact of lipedema, we performed multiple linear regression to identify predictors of quality-of-life impairment in 310 participants with complete data. The model explained 29.5% of the variance in quality-of-life scores (R² = 0.295; Adjusted R² = 0.281; F(6, 303) = 21.16, p < 0.001). As shown in Table 8, the pain scale emerged as the dominant predictor (β = 0.641; 95% CI: 0.473-0.808; p < 0.0001), with standardized coefficients indicating that pain intensity was by far the most influential factor affecting quality of life. BMI was the second significant predictor (β = 0.364; 95% CI: 0.194-0.535; p < 0.0001), suggesting that body composition independently contributes to functional limitations beyond its correlation with symptom severity. Notably, the degree of worsening with contraceptives did not emerge as a significant predictor of quality-of-life impact after adjustment for confounding factors (β = 0.106; 95% CI: -0.054 to 0.267; p = 0.1937), suggesting that hormonal effects on symptoms may influence quality of life through complementary pathways mediated primarily by pain and body composition rather than through direct independent effects.

**Table 8 -.** Multiple linear regression model for predictors of quality-of-life impact (n=310)

| Variable | $\beta$ (standardized) | 95% CI | p-value |
| --- | --- | --- | --- |
| Age | 0.061 | −0.105 to 0.226 | 0.4718 |
| BMI | 0.364* | 0.194 to 0.535 | <0.0001 |
| Symptom score | 0.044 | −0.137 to 0.225 | 0.6346 |
| Pain scale (F6) | 0.641* | 0.454 to 0.827 | <0.0001 |
| Degree of worsening with contraceptive | 0.106 | −0.054 to 0.266 | 0.1943 |
| Number of comorbidities | 0.078 | −0.085 to 0.240 | 0.348 |

Dependent variable: quality-of-life impact score (0-15 points). All continuous variables were standardized prior to analysis. Model fit: $R^2=0.295$ ; Adjusted $R^2=0.281$ ; $F(6,303)=21.16$ , $p<0.001$ . \* $p<0.05$ . $\beta$ = standardized regression coefficient; CI = confidence interval; BMI = body mass index; QoL = quality of life. Sample includes only participants with complete data on all model variables.

In summary, these findings demonstrate a significant and clinically relevant association between hormonal contraceptive use and lipedema symptom worsening in a substantial majority of users, with symptom severity, age at menarche, and specific side effects (particularly weight gain and mood changes) serving as important modulating factors. The impact on quality of life is primarily mediated through pain intensity and BMI, with chronic pain representing the most important modifiable target for therapeutic intervention. The temporal coincidence of symptom onset with contraceptive initiation in 15.1% of participants, combined with the high prevalence of perceived worsening, suggests that exogenous hormones may play an important role in both the triggering and progression of lipedema manifestations, although the cross-sectional design precludes definitive causal inference.

## DISCUSSION

This study, based on the largest Brazilian sample investigated on lipedema (n=637), provides robust and clinically relevant evidence of a significant association between hormonal contraceptive (HC) use and the worsening of lipedema symptoms. This high prevalence of perceived worsening, statistically significant (p < 0.001), corroborates previous reports in the literature [2,4,5] and transforms a frequent clinical observation into a measurable scientific finding, suggesting that exogenous hormones play an important role in the pathogenesis and progression of lipedema [1].

It is fundamental, in the analysis of these results, to differentiate direct statistical associations from broader theoretical interpretations. The data from the present study firmly support the main association: a high prevalence (58.8%) of the *perception* of lipedema symptom worsening associated with hormonal contraceptive use. Similarly, regressions robustly identify the pain scale (β=0.641) and BMI (β=0.364) as the dominant predictors of negative impact on quality of life. However, broader explanatory theories, particularly the "inflammatory model", should be treated as speculative [1,10–12]. The interpretation that the absence of correlation with duration of use (p=0.402) suggests an "individual susceptibility" instead of a cumulative effect is a working hypothesis, not a proven conclusion, since the study did not measure objective inflammatory or lymphatic biomarkers. This clear distinction between statistical association and causal hypothesis is crucial for the correct interpretation of the findings.

These findings reinforce the hypothesis that lipedema is not only modulatable by hormonal factors but also deeply influenced by inflammatory and lymphatic pathways, in which contraceptives act as contextual amplifiers.

### Evidence of a Possible Causal Relationship

An interesting finding was that 15.1% of participants (n=96) reported that the onset of lipedema symptoms coincided temporally with the start of contraceptive use. Although this finding suggests a possible temporal association—one of the Bradford Hill [13] criteria for causality—it is important to emphasize that in cross-sectional studies it is not possible to distinguish between: (a) contraceptives as a triggering factor; (b) unmasking of pre-existing subclinical disease; or (c) recall bias. Prospective studies are necessary to clarify this relationship. When combined with the strong dose-response association observed between symptom score and risk of worsening (OR = 1.562; 95% CI: 1.300–1.877; p < 0.001), where each additional symptom increased the risk of worsening by 50%, the hypothesis of a relationship possibly mediated by plausible biological mechanisms is strengthened [14,15], even without direct causal confirmation.

### Pathophysiological Mechanisms and Key Intermediaries

A relevant finding was that the duration of contraceptive use did not predict worsening (OR 0.931; p=0.402), nor did age (OR 0.926; p=0.395), indicating that the adverse response does not follow a cumulative dose-dependent pattern. Significant predictors were baseline symptom score (OR 1.562; p<0.001), suggesting that pre-existing severity determines vulnerability to hormones, while age at menarche (OR 0.746; p=0.014) and BMI (OR 0.819; p=0.037) showed modest protective effects, possibly reflecting differences in metabolic-hormonal programming. A possible interpretation is that worsening with contraceptives may reflect individual susceptibility rather than a dose-dependent effect, where predisposed women manifest worsening early while non-susceptible ones tolerate prolonged use. Alternatively, this finding may reflect survival bias (women who worsened significantly discontinued use early) or insufficient statistical power to detect weak associations. The fact that baseline symptom score was the strongest predictor (OR 1.562; p<0.001) suggests that the pre-existing severity of the condition may influence the response to exogenous hormones, although the exact mechanism remains to be elucidated. This explains why the strongest predictor was precisely the *baseline symptom score* (OR 1.562; p<0.001)—a direct marker of pre-existing inflammatory activity—and not the duration of exposure. Age at menarche (OR 0.746; p=0.0135), in turn, acts as a temporal proxy for immunometabolic programming: early menarche implies a larger window of inflammatory "imprinting" during tissue maturation, establishing elevated baseline reactivity. In summary, lipedema associated with contraceptives seems to emerge from *constitutive vulnerability* amplified by hormones, not toxic accumulation—a paradigm that reorients clinical focus from duration of use to the early identification of risk phenotypes.

A counter-intuitive finding deserves highlighting: the duration of contraceptive use was not a significant predictor of worsening in the multivariate analysis (OR 0.931, p=0.4018). If worsening were a cumulative dose-dependent phenomenon, we would expect longer duration of use to correlate with a higher probability of deterioration. The absence of this relationship suggests an alternative mechanism: instead of a cumulative effect, there seems to be individual susceptibility, where vulnerable women manifest worsening early (possibly in the first few cycles), while non-susceptible ones may use contraceptives indefinitely without significant deterioration.

The analysis of side effects revealed a strong association between weight gain and lipedema worsening (χ² = 29.32, p < 0.0001): 71.7% of women who experienced weight gain reported symptom worsening, compared to 43.5% of those without weight gain—a difference of 28.2 percentage points. This finding admits multiple interpretations. First, metabolic and fluid retention effects of contraceptives may exacerbate pre-existing lipedema symptoms, creating an overlap between iatrogenic side effects (hormone-induced edema) and manifestations of the underlying disease [16–19]. Second, weight gain may reflect a marker of individual hormonal response, where women more metabolically sensitive to contraceptives also present greater reactivity in the adipose tissue affected by lipedema [20–22]. Third, reverse causality cannot be excluded, where lipedema worsening (with increased edema and tissue volume) is interpreted by participants as "weight gain". The cross-sectional design of the study does not allow distinguishing between these hypotheses, although all are biologically plausible and not mutually exclusive.

Mood alteration as a side effect also showed a significant association with lipedema worsening (77.4% vs 51.3%, p = 0.0002). This finding can be interpreted in different ways. One possibility is that mood changes reflect increased sensitivity to the systemic effects of exogenous hormones, potentially indicating greater individual susceptibility both in the central nervous system and in other target tissues, including adipose tissue [23,24].

Alternatively, women with more severe lipedema symptoms may present a higher baseline psychological stress load, making them more vulnerable to contraceptive-induced mood changes—representing confounding rather than direct causality [3]. Finally, both mood symptoms and lipedema worsening may be influenced by unmeasured variables (such as stress, sleep, dietary factors), creating a spurious association. Prospective studies with standardized psychometric assessment and biological markers of hormonal sensitivity are necessary to elucidate the nature of this association.

### Interpretative Framework: Inflammatory Model of Lipedema

The magnitude of the observed association is clinically significant: among 588 hormonal contraceptive users, 58.8% reported symptom worsening (χ²=213.71; p<0.001), a distribution that differs significantly from random expectation. Particularly relevant, 15.1% of participants (n=96) reported temporal coincidence between the start of contraceptives and symptom onset, suggesting a possible temporal association that merits prospective investigation.

#### Integration with Pathophysiological Models Proposed in the Literature

Although the present study did not include objective measures of inflammation or lymphatic assessment, the findings are remarkably consistent with emerging pathophysiological models in the literature that propose lipedema as a chronic inflammatory syndrome of soft tissues. Several authors have suggested an integrated inflammation → lymphatic → adipose model, in which low-grade inflammatory processes precede and perpetuate the characteristic changes of the disease [25,26].

In this conceptual framework, mast cell activation, vascular permeability alterations, progressive lymphatic dysfunction, and the release of inflammatory mediators (histamine, tryptase, prostaglandins, cytokines) would create a microenvironment conducive to disproportionate adipocyte hyperplasia and fibrosis [26–31]. Recent multi-omics studies have identified characteristic molecular signatures, including polarized M2 macrophages, elevated CD163 expression, interstitial hypoxia, and extracellular matrix remodeling mediated by metalloproteinases [11,12,32].

These findings refine our understanding of the ’Immunological Shield,’ suggesting that the protection conferred by lipedema tissue comes at a homeostatic cost. We know from the ’Menopausal Switch’ concept that estrogen is vital for maintaining the ’Metabolic Sink’s’ protective function against cancer. However, the introduction of synthetic hormones appears to create a paradox: they maintain the ’systemic protection’ signal but overwhelm the local vascular and lymphatic drainage capacity. Therefore, the hormone is not the villain, but a potent modulator that, in a predisposed phenotype, can tip the balance between systemic immunometabolic advantage and local symptomatic comfort, necessitating a personalized clinical approach rather than the demonization of female endocrinology [12,32–34].

#### Connection between Study Findings and the Inflammatory Model

Several findings from the present study, while not proving this model, are consistent with its predictions:

**First**, the absence of an effect of duration of use (OR 0.931; p=0.4018) contradicts the cumulative hormonal toxicity model [35–37] and suggests individual susceptibility— consistent with the hypothesis that women with already elevated tissue "inflammatory tone" respond adversely from the beginning of exposure, regardless of duration.

**Second**, the baseline symptom score as the dominant predictor of worsening (OR 1.562; p<0.001) suggests that pre-existing disease activity—a possible indirect marker of underlying inflammatory process—determines vulnerability to exogenous hormones.

**Third**, the protective association of age at menarche (OR 0.746; p=0.0135) could reflect differences in inflammatory "programming" during critical developmental windows, although alternative interpretations (confounding, chance) cannot be excluded [38–41]. The hypothesis would be that later menarche implies less cumulative exposure to estrogenic fluctuations during the maturation of immune and vascular systems in subcutaneous adipose tissues.

**Fourth**, the most prevalent side effects—swelling (40.5%) and weight gain (41.8%)— are expected manifestations of both inflammatory fluid retention and direct hormonal effects, making the distinction between cause and consequence difficult [16–19,42,43].

#### Hormones as Modulators, Not Causers

This framework suggests that sex hormones— endogenous or exogenous—would not be the primary cause of lipedema, but gain modulators that amplify pre-existing inflammatory vulnerability. This would explain why milestones of intense hormonal variation (puberty, pregnancy, contraceptives, menopause) frequently coincide with symptom onset or exacerbation, without being direct causers [3,44]. Estrogens and progestogens would modulate mast cells, endothelium, and lymphatic function, amplifying circuits already predisposed to dysfunction [45–47].

From an evolutionary medicine perspective, it is speculated that lipedema may represent a maladaptive response of ancestral protective peripheral reserve programs when confronted with modern environments characterized by chronic inflammatory exposure (pro-inflammatory diet, sedentary lifestyle, environmental endocrine disruptors, sleep deprivation). In this context, continuous exogenous hormonal exposure would act as an amplifier of an already inflammo-sensitive phenotype [48].

#### Critical Limitations and Need for Validation

It is imperative to emphasize that these interpretations are speculative and based on literature synthesis, not direct measures. The present study:

Did not measure inflammatory biomarkers (CRP, IL-6, TNF-α, tryptase). Did not objectively assess lymphatic function (lymphoscintigraphy, ICG). Did not perform histopathological or molecular analysis.

Cannot distinguish between genuine pathophysiological worsening of lipedema versus independent side effects (iatrogenic edema) interpreted as disease worsening.

Therefore, the inflammatory model remains an attractive and biologically plausible hypothesis, but not validated by the data of the present study.

### Immediate Clinical Implications

The findings not only elucidate etiological aspects but also have direct implications for clinical practice and the design of therapeutic interventions. Before starting contraceptives, clinicians should actively investigate signs of lipedema using validated screening tools integrated into routine gynecological protocols. Women with confirmed or suspected lipedema should receive explicit information about the risk of worsening to allow for an informed choice between hormonal contraceptives and non-hormonal methods. In women who experience worsening, evidence-based adjuvant measures for chronic inflammatory conditions may be considered, including nutritional optimization, exercises with lymphatic emphasis, and lifestyle factor management. Furthermore, longitudinal research testing inflammatory reversibility and pragmatic trials comparing strategies centered on inflammatory control are necessary. Finally, early and objective monitoring is recommended for women who choose to continue contraceptives despite the risk

### Considerations on Pharmacological Safety

Although this study did not segregate results by progestogen type, pharmacological literature imposes specific caution regarding the use of drospirenone in patients with Lipedema. Robust evidence, including large cohorts and meta-analyses, demonstrates that combined oral contraceptives containing drospirenone are associated with a venous thromboembolism (VTE) risk 1.5 to 3 times higher compared to second-generation contraceptives containing levonorgestrel levonorgestrel [49–51]. Considering that women with Lipedema frequently present elevated baseline risk factors—such as increased BMI, venous stasis, or low-grade chronic inflammation—the addition of an agent with greater thrombogenic potential constitutes a dangerous and unnecessary risk association [3,27,52]. Therefore, we do not recommend the prescription of drospirenone for this population; when hormonal use is imperative, formulations with a safer hemostatic profile (such as levonorgestrel) or non-hormonal methods should be prioritized.

### Functional and Psychosocial Impact

The impairment of quality of life was evident and quantifiable in the studied sample. Multivariate linear regression analysis identified that the main independent predictors of greater impact on quality of life were the pain scale (β = 0.641; p < 0.001) and BMI (β = 0.364; p < 0.001). Notably, the degree of worsening with contraceptives did not emerge as a significant predictor after adjustment for confounding factors (β = 0.106; p = 0.194), suggesting that the impact on quality of life is primarily mediated by pain intensity and body composition, regardless of the perception of hormonal worsening.

The psychosocial impact was equally striking, with 70.5% of participants reporting low or fair self-esteem, and a median pain score of 5 on a 0-10 scale. These data highlight the double burden imposed by lipedema—physical and emotional—which may be exacerbated by hormonal contraceptive use, reinforcing the need for a comprehensive approach in the management of these patients [3,53,54].

### Methodological Innovation and Scientific Contribution

An important methodological differentiator of this study was the innovative use of natural language processing via local LLM to systematically categorize 19 free-text variables, including side effects, treatments, comorbidities, and timing of symptom onset. This approach allowed the quantification of qualitative data that would traditionally be lost in conventional statistical analyses or require intensive manual categorization, representing a significant methodological advance for lipedema research. The precise identification that 15.1% of women associated symptom onset specifically with HC use was made possible by this technique, highlighting its value in capturing important clinical nuances.

### Limitations and Future Directions

From an evolutionary perspective, lipedema may represent the modern maladaptation of an ancient protective peripheral reserve program that, in environments of inflammatory ultra- exposure (pro-inflammatory diet, dysbiosis and metabolic endotoxemia, environmental endocrine disruptors, sleep deprivation/chronodisruption, sedentary lifestyle), remains chronically activated [48]. Environmental xenoestrogens, such as bisphenol A and phthalates, act as co-factors, mimicking estrogen and exacerbating endothelial and mast cell dysfunction [55–57]. Continuous exogenous hormonal exposure acts as a modulator that exacerbates this inflammatory-lymphatic phenotype, not necessarily as its initiator. In this frame, lipedema emerges as a phenotype of programmed inflammatory vulnerability, upon which hormones operate as contextual amplifiers. Resistance to muscle mass gain and weight loss through exercise—frequently mentioned by patients—reflects that the tissue is not metabolically normal fat, but an inflamed and fibrotic matrix resistant to conventional lipolysis [58–60].

If this model is confirmed in future studies with objective measures, it would have important therapeutic implications:

**Pre-prescription screening:** Identify women with elevated inflammatory phenotype (hs-CRP, baseline tryptase) before starting contraceptives.

**Multimodal anti-inflammatory interventions:** Test strategies combining anti-inflammatory nutrition, mast cell stabilizers (ketotifen, cromolyn), targeted supplementation (curcumin, quercetin, PEA), sleep optimization, and lymphatic exercise [61–67].

**Objective monitoring:** Track biomarkers before and after interventions to assess reversibility.

**Risk stratification:** Develop predictive scores integrating baseline symptoms, hormonal history, and inflammatory markers.

This study presents limitations that must be considered. First, the cross-sectional design does not allow establishing definitive causality, only temporal association. Second, data are self-reported, without objective validation by anthropometric measures, inflammatory biomarkers, or imaging exams—making it impossible to categorically distinguish between genuine pathophysiological worsening of lipedema versus independent side effects (iatrogenic fluid retention) interpreted as worsening of the underlying disease. Third, recall bias may affect memory regarding timing of symptom onset and medication use. Fourth, a convenience sample recruited from support groups may overestimate the association (selection bias). Finally, it was not possible to stratify by specific type of contraceptive, dosage, or route of administration. Despite these limitations, the large sample size (n=637), robust multivariate analyses, and magnitude of the observed effect (58.8%) confer substantial validity to the findings.

It is worth acknowledging an important methodological limitation: without objective measures (inflammatory biomarkers, lymphoscintigraphy, soft tissue ultrasonography), it is not possible to categorically distinguish between (A) genuine pathophysiological worsening of lipedema, (B) independent side effects of contraceptives (iatrogenic fluid retention) erroneously interpreted as worsening of the underlying disease, and (C) hypervigilance bias in women with already established severe symptoms. Future studies should incorporate objective endpoints—variation in standardized circumferences, body water index by bioimpedance, serum tryptase dosage, and vascular permeability markers—to separate real inflammatory signal from perceptual noise. This caveat, however, does not invalidate the observed association pattern, only nuances its mechanistic interpretation. Still, the consistency of findings across multiple analyses, the temporal coherence, and the magnitude of observed effects confer epidemiological credibility to the identified pattern.

Prospective studies should follow women before the start of contraceptives with objective measures (circumferences, bioimpedance, inflammatory biomarkers) to establish temporal causality. Controlled interruption trials are needed to test symptom reversibility after hormonal discontinuation, ideally also comparing multimodal anti-inflammatory interventions in a factorial design. Molecular characterization via subcutaneous tissue biopsy and pharmacogenomic studies could identify susceptibility phenotypes and precise therapeutic targets. Finally, stratification by type, dose, and route of administration of contraceptives would allow identifying hormonal profiles with less deleterious impact in women with lipedema.

## CONCLUSION

In summary, this study demonstrated a significant and clinically relevant association between hormonal contraceptive use and the worsening of lipedema symptoms. The findings are consistent with the hypothesis that exogenous hormones may modulate the clinical expression of the disease in predisposed women. Although the exact mechanisms remain to be elucidated, the data suggest that baseline symptom score and reproductive history may influence individual response to contraceptives. These findings have immediate implications for contraceptive counseling of women with lipedema, while prospective studies with objective measures are needed to establish causality and mechanisms.

## Data Availability

All data produced in the present study are available upon reasonable request to the authors

## APPENDIX

### Appendix Table 1. Study Questionnaire

| Section | Question | Type / Options |
| --- | --- | --- |
| **A. Consent & ID** | A1. Consent | Checkbox (Required) |
|  | A2. Age | Number (18-100) |
|  | A2.1 Current Weight (kg) | Decimal Number |
|  | A2.2 Height (cm) | Number |
|  | A2.3 State of Residence | Dropdown (BR States) |
| **B. Screening** | B1. Do you feel pain in your legs? | Yes / No / Sometimes |
|  | B2. Do your legs swell during the day? | Yes / No / Sometimes |
|  | B3. Do symptoms worsen with heat? | Yes / No |
|  | B4. Are your legs sensitive to touch? | Yes / No |
|  | B5. Do you bruise easily on your legs? | Yes / No |
|  | B6. Do you notice disproportionate fat accumulation? | Yes / No |
|  | B7. Do diets and exercise fail to reduce leg size? | Yes / No |
|  | B8. Is there a family history of lipedema? | Yes / No / Don't know |
|  | B9. When did you first notice these symptoms? | Puberty / Pregnancy / Menopause / Contraceptive use / Other |
| **C. Diagnosis** | C1. Have you been diagnosed with lipedema? | Yes / No / Suspected |
|  | C2. Diagnosis Date | Month/Year |
|  | C4. Lipedema Stage | Stages 1-4 / Don't know |
|  | C5. Lipedema Type | Types I-V / Don't know |
|  | C6. Other medical conditions | Long Text |
| **D. Hormonal** | D1. Age at menarche | Number |
|  | D2. Menstrual cycle regularity | Yes / No / Menopause |
|  | D4. Number of pregnancies | Number |
|  | D6. Menopause status | Yes / No / Perimenopause |
|  | D9. Thyroid alterations | Hypo / Hyper / No |
| **E. Contraceptives** | E1. Do you use/have used hormonal contraceptives? | Current / Previous / Never |
|  | E2. Type of contraceptive | Pill / Injection / IUD / Implant / Patch / Ring |
|  | E3. Brand Name | Text |
|  | E4. Duration of use | Text |
|  | E5. Age at start of use | Number |
|  | E6. Pattern of use | Continuous / Interrupted |
|  | E9. Side effects noticed | Weight gain / Swelling / Headache / Mood / etc. |
|  | E10. Worsening of lipedema after starting? | Severe / A little / None / Improved |
|  | E12. Weight gain after starting? | Yes / No |
| **F. Quality of Life** | F1. Impact on mobility | Very much / Moderately / Little / None |
|  | F3. Impact on work | Yes / No / Don't work |
|  | F6. Average daily pain (0-10) | Number (0-10) |
|  | F8. Self-esteem | Excellent / Good / Fair / Low |

---

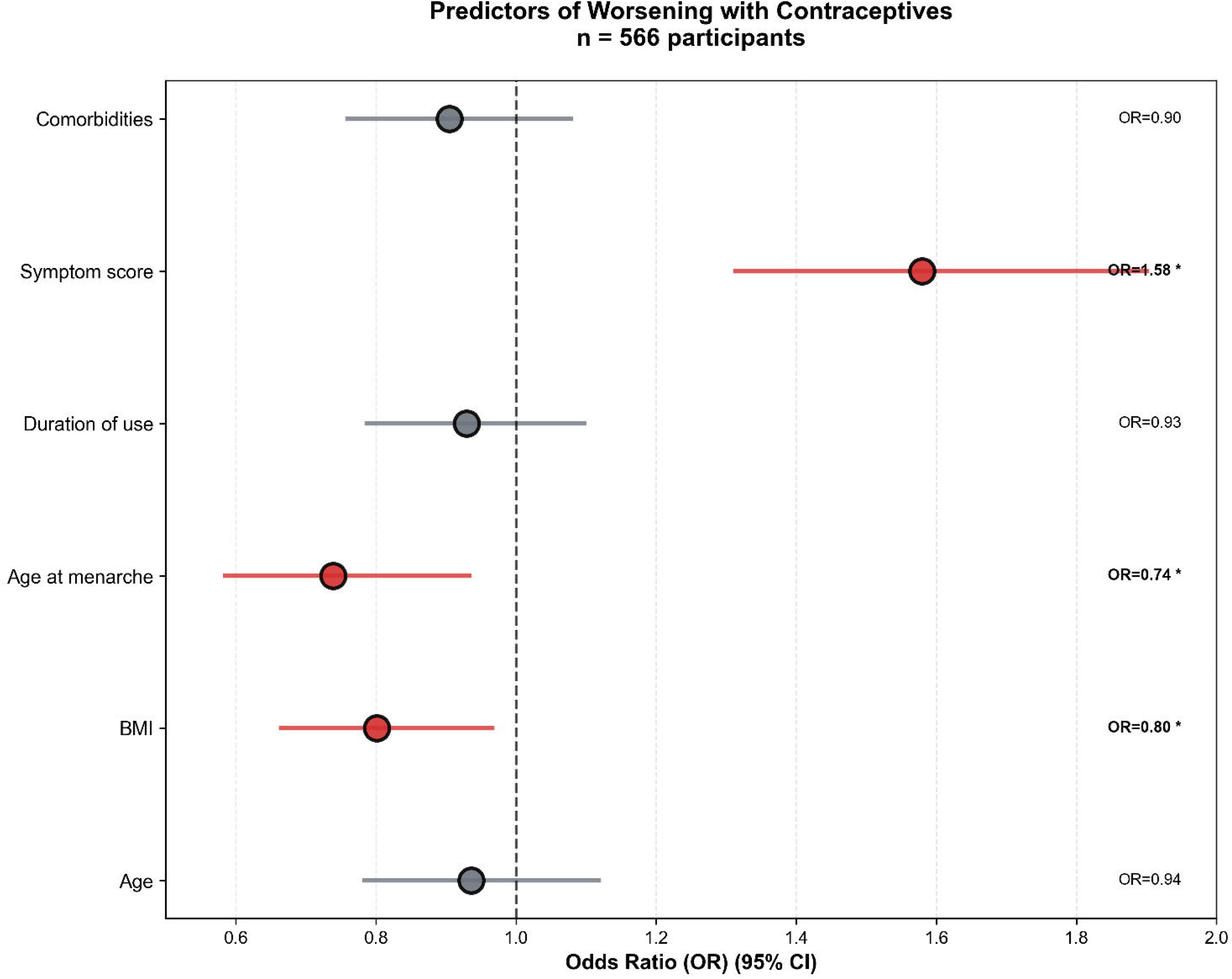

## Notes

### Competing Interest Statement

The authors have declared no competing interest.

### Funding Statement

This study did not receive any funding

